# Effects of the introduction of interprofessional conferences on the intensive care unit: Comparison of the length of stay in the intensive care unit before and after the introduction of interprofessional conferences

**DOI:** 10.1101/2022.04.24.22273777

**Authors:** Daichi Watanabe, Keiichi Uranaka, Kyouko Asazawa, Takako Akimoto, Hironori Ohnuma

## Abstract

Recently, interprofessional team medicine has been practiced to achieve medical safety and improve patient outcomes. This study evaluated the effects of interprofessional conferences on intensive care units (ICUs) by comparing outcomes before and after their introduction. This study was conducted at a single center and included 1,765 patients who were admitted to the ICU between April 2017 and March 2019. There were 898 patients in the group before the introduction of conferences (before group) and 867 patients in the group after the introduction of conferences (after group). The interprofessional conferences involved physicians, nurses, physical therapists, nutritionists, and pharmacists. Data were extracted from the medical records. The primary outcome measure was the ICU length of stay (LOS). The secondary outcome measures were hospital LOS and rehabilitation and nutrition started within 48 hours of ICU admission. These outcomes were compared before and after the introduction of interprofessional conferences. The adjusted variables were gender, age, body mass index, ICU readmission, outcome, Barthel index at admission, and disease (classified by the International Statistical Classification of Diseases and Related Health Problems 10^th^ edition). The ICU LOS (regression coefficient: -0.08; 95% confidence interval [CI]: -0.13 to -0.04) and hospital LOS (regression coefficient: -2.96; 95% CI: -5.20 to -0.72) were significantly shorter in after group than in the before group. Moreover, the proportion of patients who commenced nutrition (odds ratio [OR]: 1.45; 95% CI: 1.14 to 1.84) and rehabilitation (OR: 0.77; 95% CI: 0.51 to 1.17) within 48 hours of ICU admission was significantly higher in the after group than in the before group. Interprofessional conferences effectively reduced the ICU LOS and hospital LOS and improved the likelihood of commencing nutrition and rehabilitation within 48 hours of ICU admission.

## Introduction

Recently, a wide variety of staff members have been practicing interprofessional team medicine based on their high levels of expertise, sharing objectives and information, and following the individualized needs of each patient [1]. In complex medical environments such as intensive care units (ICUs), where critically ill patients with limited physiological reserves undergo several tests and treatments, effective communication among professions is especially important as part of team medicine [2]. In terms of medical safety, according to the Joint Commission on Accreditation of Health Organization, communication errors are a major cause of medical errors, which comprise 85% of serious medical incidents in hospitals. It has been suggested that effective communication can reduce medical errors and improve short-term patient outcomes such as mortality and readmissions [3]. Additionally, the United States Critical Illness and Injury Trial Group Critical Illness Outcomes Study has identified that the establishment of a daily medical plan and a lower nursing ratio are factors that improve patient outcomes [4]. Therefore, the management of critically ill patients by an interprofessional team led by an intensivist has been proposed to improve the quality of care by strengthening interprofessional collaboration, with all professionals having parts of the roles performed by intensivists and utilizing their own expertise [5]. To provide high-quality patient care, it is essential to share patient care plans among many professionals and facilitate communication among them.

Previous studies performed in Japan and other countriess have evaluated the introduction and implementation of interprofessional protocols, interprofessional rounds, and interprofessional conferences and their effects on ICU outcomes. As an example of interprofessional protocols, for patients requiring ventilation for more than 48 hours in the ICU, an interprofessional team examined the effects of shallow sedation management on safety and ventilator duration using a quality improvement model. The results of this study reported shallower sedation according to the Richmond Agitation-Sedation Scale during the day, decreased unplanned device extractions, and decreased ventilator-related adverse events [6]. Additionally, a study of the effects of introducing a long-term ventilator weaning plan led by an interprofessional team for patients requiring ventilation in the ICU for more than 21 days reported decreased ICU and in-hospital mortality rates and decreased ventilator durations [7]. In terms of nutrition, the effects of interprofessional collaboration have also been evaluated. One study reported that the introduction of an interprofessional ICU nutrition management protocol resulted in an improvement in the early enteral nutrition rate within 48 hours of admission to the ICU [8]. Therefore, the introduction of interprofessional protocols is associated with improved patient outcomes in terms of sedation, ventilator use, and nutrition.

Furthermore, a study of the effects of daily bedside interprofessional rounds on the outcomes of patients admitted to ICUs at 112 facilities found that interprofessional rounds were associated with decreased mortality rates within 30 days of admission [9]. Additionally, one study of the effects of the participation of family members of ICU patients in interprofessional rounds showed that this participation was associated with better family satisfaction in terns of frequency of communication with the physician and support during decision-making [10]. Therefore, interprofessional rounds contribute not only to better patient outcomes but also to better patient and family satisfaction rates.

Moreover, one study of the effects of introducing interprofessional conferences in an open-system ICU reported that the number of days before rehabilitation commenced was reduced after their introduction compared to the number before their introduction [11]. Another study [12] verified the beneficial effects of introducing interprofessional conferences on rehabilitation, including the treatment plan by cardiovascular surgeons and physical therapists, in the ICU, and it reported decreased endotracheal intubation durations, decreased mean hospital length of stay (LOS), and decreased mean ICU LOS compared to those before the introduction of interprofessional conferences. However, all of these studies of interprofessional conferences involved descriptive statistics or univariate analyses, and confounding factors were not adjusted for, thus making it impossible to conclude the effectiveness of interprofessional conferences. Therefore, we evaluated the effects of interprofessional conferences that were introduced for the purpose of sharing treatment plans and information and making decisions about care and medicine to be provided as part of interprofessional team medicine by performing a multivariate analysis to adjust for confounding factors and using patient outcomes as indicators.

## Materials and Methods

### Terms definition

During this study, an interprofessional conference was defined as an interprofessional team consisting of a physician and at least one other health care provider [3] sharing treatment plans and information and making decisions about the care to be provided in the ICU.

### Conceptual framework

The conceptual framework of this study is shown in (Fig 1). During this study, the effects of interprofessional conferences in ICUs were evaluated before and after their introduction. The primary outcome measure was the ICU LOS. The ICU LOS before the introduction of interprofessional conferences and that after their introduction were compared. Secondary outcome measures were hospital LOS, whether nutrition was started within 48 hours of ICU admission, whether rehabilitation was started within 48 hours of ICU admission, and mortality within 28 days of ICU admission. The adjusted variables were gender, age, body mass index (BMI), disease (International Statistical Classification of Diseases and Related Health Problems 10^th^ edition [ICD-10]), Barthel index (BI) at admission, outcome, ICU readmission, and surgical procedure.

**Figure 1:**
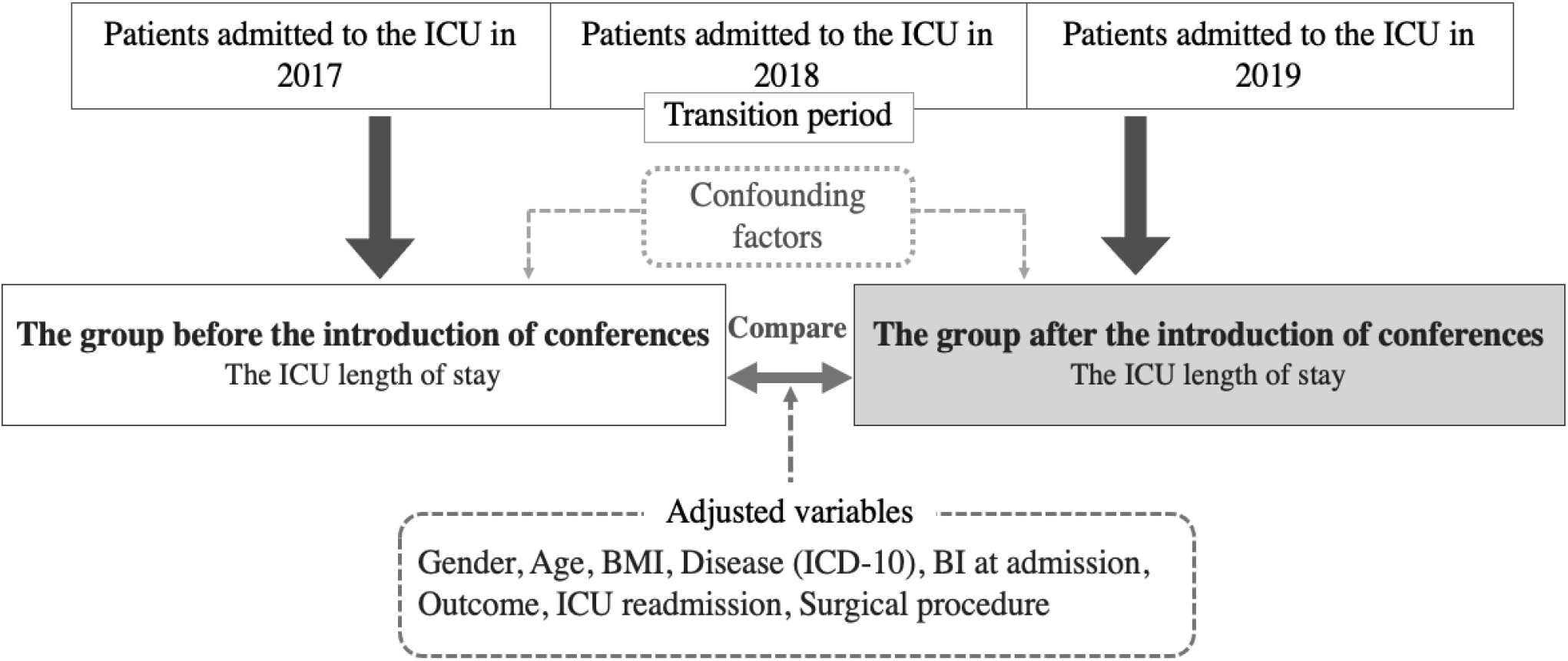
The conceptual framework of this study BMI: Body mass index, ICD-10: International Statistical Classification of Diseases and Related Health Problems 10^th^ edition, BI: Barthel index

### Design

This was a retrospective observational study. It included medical records from April 2017 to March 2019.

### Study population and setting

The inclusion criterion was admission to the ICU during April 2017 to March 2019. The exclusion criteria were admission to the ICU for less than one night and age 18 years or younger. A specific ICU at a municipal hospital with approximately 700 beds in central Hokkaido, Japan, was selected as the research facility for the study. The ICU at the study site was a medical/surgical ICU with 16 beds and was operated using a semi-closed system. It was staffed by an intensivist and had a patient-to-nurse ratio of 2:1. Staff members during the day shift included a full-time physical therapist, clinical engineer, nutritionist, and pharmacist. The research facility introduced regular interprofessional conferences as part of interprofessional team care in 2018. In 2019, an operation manual for interprofessional conferences was created within the ICU department, and the format for recording the conferences in the electronic medical records was standardized. Conferences were conducted once per week for approximately 30 minutes. One target patient was determined after a discussion among the interprofessional staff the day before the conference. There was no clear definition of eligible patients; however, priority was given to patients who were critically ill, patients whose care or treatment was not progressing as planned, and patients who were at high risk for such problems.

The interprofessional conference participants were physicians, nurses, physical therapists, nutritionists, pharmacists, clinical engineers, and medical social workers. The purpose of the interprofessional conferences and the contents of discussions varied depending on the target patient; however, the main objectives were sharing treatment plans and information and making decisions about the care and treatment to be provided. Additionally, patient goals and their evaluation dates, as well as specific care plans to achieve them, were discussed.

### Data collection

After obtaining consent for research cooperation from the administrators of the collaborating facilities, patient data and the number of conferences conducted were extracted from the medical records for the study. The survey items included the following patient characteristics: gender; age; height; weight; disease name (ICD-10); patient outcome (alive or dead); BI at admission (calculated using a 20-point scale for this study); surgical procedure (including scheduled and emergency surgeries); ICU readmission; ICU admission date; ICU discharge date; hospital admission date; discharge date; whether nutrition was started within 48 hours (including enteral and oral nutrition); whether rehabilitation was started within 48 hours of ICU admission (rehabilitation was defined as intervention by a physical therapist); and mortality within 28 days of ICU admission.

### Measurements

#### Intensive care unit length of stay

The ICU LOS was calculated as the difference in the number of days between the date of admission to the ICU and the date of discharge from the ICU. If the patients were re-admitted to the ICU within the same admission period, then they were treated as the same patient, and the LOS in the ICU was calculated by adding the number of days spent in the ICU after the first and subsequent ICU admissions.

#### Hospital length of stay

The hospital LOS was calculated as the number of days between admission to the study facility and the date of discharge from the facility. If the study patients were discharged and readmitted, then they were treated as different patients and their hospital LOS were calculated separately.

#### Other variables related to the intensive care unit length of stay

Factors related to the ICU LOS were based on previous studies and included patient age, gender, outcome, surgical procedure, and disease [11]. The BMI [6], ICU readmission [13,14], and BI at admission [15,16] were also selected as variables.

### Statistical analysis

Descriptive statistics were calculated for each variable, and patient characteristics before and after the introduction of the interprofessional conferences were compared using the Student t test and χ^2^ test, depending on the type of variable. The Mann-Whitney U test was also used to compare the median number of days in the ICU for each variable before and after the introduction of interprofessional conferences. Correlations between variables were calculated using Pearson’s correlation coefficient and the Spearman rank correlation coefficient, depending on the type of scale. To examine differences in the ICU LOS based on the introduction of interprofessional conferences, the dependent variable was ICU LOS and the independent variables were before and after the introduction of interprofessional conferences, gender, age, BMI, ICU readmission, outcome, BI at admission, and disease (ICD-10); these were forced variables and a Poisson regression analysis was performed. A multiple regression analysis was performed for the secondary outcome measures; the dependent variable was hospital LOS and the independent variables were before and after the introduction of interprofessional conferences, gender, age, BMI, ICU readmission, outcome, BI at admission, and disease (ICD-10). Regarding whether nutrition was started within 48 hours of ICU admission and whether rehabilitation was started within 48 hours of ICU admission, the dependent variables were whether nutrition was started within 48 hours of ICU admission and whether rehabilitation was started within 48 hours of ICU admission, respectively; the independent variables were before and after the introduction of interprofessional conference, gender, age, BMI, ICU readmission, outcome, BI at admission, and disease (ICD-10). We forced those independent variables and performed a logistic regression analysis. Regarding mortality within 28 days of ICU admission, a logistic regression analysis was performed with mortality within 28 days of ICU admission as the dependent variable and forced entry of before and after the introduction of interprofessional conferences, gender, age, BMI, ICU readmission, BI at admission, and disease (ICD-10) as the independent variables.

All statistical analyses were performed using Stata version 15 (Stata Corp LLC, Lakeway, TX, USA). Two-sided p<0.05 was considered statistically significant.

### Ethical considerations

Ethical approval for this study was obtained from the Ethics Review Committee of Teine Keijinkai Hospital (no. 2-020276-00) and from the Ethics Committee of Tokyo Healthcare University of the researcher (no. 32-50). Informed consent was obtained using a form provided on the website of the Teine Keijinkai Hospital; ethical approval was given. There are no conflicts of interest to disclose.

## Results

Details of the interprofessional conferences conducted are shown in Table 1. There were 58 interprofessional conferences conducted in 2019, and 22 patients were the target of those conferences (in some cases, more than one conference was conducted for a single patient). The contents of 31 conferences included sharing information about the patients. The contents of 25 conferences included making clear decisions about treatment and treatment plans. The contents of two conferences included ethics. There were no conferences about unclear records or unavailable records.

**Table1.**
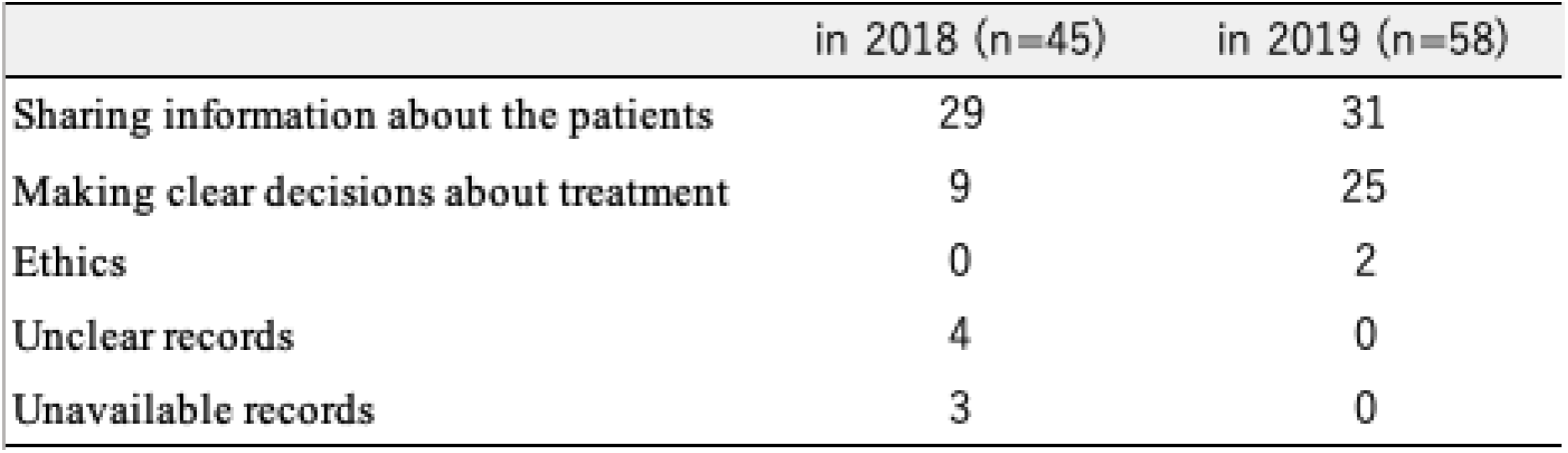
Number and contents of interprofessional conference

|  | in 2018 (n=45) | in 2019 (n=58) |
| --- | --- | --- |
| Sharing information about the patients | 29 | 31 |
| Making clear decisions about treatment | 9 | 25 |
| Ethics | 0 | 2 |
| Unclear records | 4 | 0 |
| Unavailable records | 3 | 0 |

The characteristics of the study participants are shown in Table 2. The number of study participants was 1765; there were 898 patients in the group before the introduction of interprofessional conferences (before group) and 867 patients in the group after the introduction of interprofessional conference (after group). The mean age of the patients was 69.7 years (standard deviation [SD]: ±13.6 years); 59% were male patients (1043) and 41% were female patients (722). The mean BMI was 23.3 (SD: 4.5). There were significantly more patients who required readmission in the before group than in the after group (χ^2^=4.76; degrees of freedom=1.0; p=0.029). There were no significant differences in other characteristics of the two groups.

**Table2.** The characteristics of the study participants (N=1765)

| Variables | Bfore group<br>(n=898) |  | After group<br>(n=867) |  | *p-value* |
| --- | --- | --- | --- | --- | --- |
|  | Mean | SD | Mean | SD |  |
| Age | 69.6 | ± 13.5 | 69.9 | ± 13.6 | 0.710 |
| BMI | 23.2 | ± 4.7 | 23.4 | ± 4.2 | 0.420 |
| BI at admission | 14.3 | ± 7.9 | 14.1 | ± 8.2 | 0.690 |
|  | n | % | n | % |  |
| Gender |  |  |  |  | 0.890 |
| Male | 532 | 51.0 | 511 | 49.0 |  |
| Female | 366 | 50.7 | 356 | 49.3 |  |
| Surgical procedure | 724 | 51.8 | 672 | 48.2 | 0.100 |
| ICU readmission | 57 | 61.9 | 35 | 38.1 | 0.029 |
| Disease (ICD-10) |  |  |  |  | 0.260 |
| Cardiovascular disease | 342 | 48.4 | 364 | 51.6 |  |
| Neoplasms | 318 | 52.3 | 289 | 47.7 |  |
| Gastrointestinal diseases | 82 | 49.1 | 85 | 50.9 |  |
| Other diseases | 155 | 54.5 | 129 | 45.5 |  |
| Patient outcome |  |  |  |  | 0.940 |
| Alive | 819 | 50.9 | 786 | 49.1 |  |
| Dead | 79 | 49.3 | 81 | 50.7 |  |

Student t test and $\chi^2$ test were used to compare patient characteristics before and after the introduction of the interprofessional conferences
Before group : the group before the introduction of conferences, After group : the group after the introduction of conferences
SD : standard deviation, BMI : Body Mass Index, BI : Barthel Index(calculated using a 20-point scale for this study)
ICD-10 : International Statistical Classification of Diseases and Related Health Problems 10th edition

Table 3 shows the results of a comparison of the before and after groups in terms of the ICU LOS and patient characteristics. There were no significant differences in the ICU LOS based on the characteristics of patients in the before and after groups.

**Table3.**
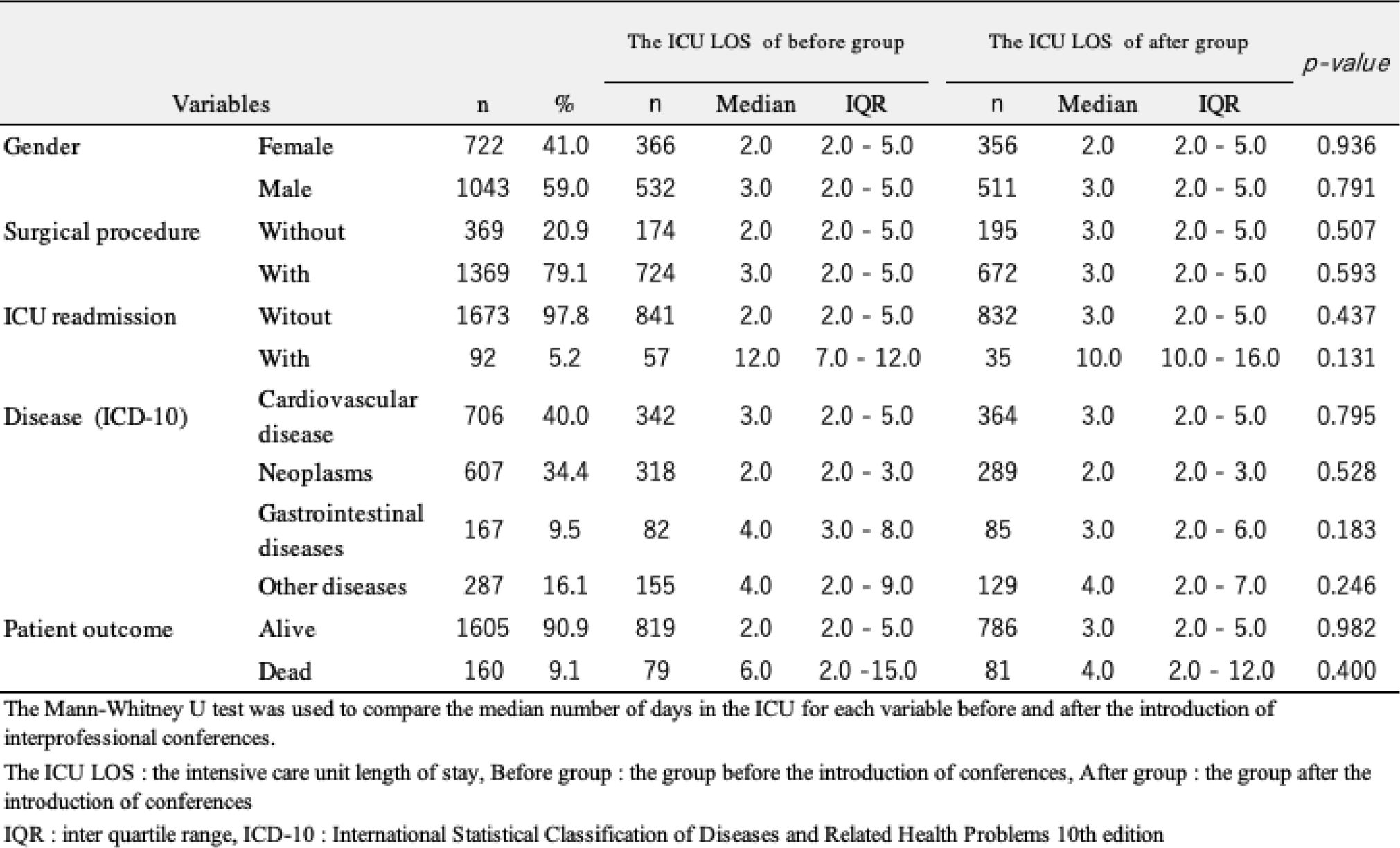
The ICU LOS and patient characteristics of the before and after groups. (N=1765)

The primary and secondary outcome measures were compared by performing a univariate analysis of the before and after groups (Table 4). The results showed no significant difference in the ICU LOS of the before and after groups (p=0.859). The proportion of patients who started nutrition within 48 hours of ICU admission was significantly higher in the after group (67.8%) than in the before group (61.9%) (p=0.009). The proportion of patients who started rehabilitation within 48 hours of ICU admission was also significantly higher in the after group (68.0%) than in the before group (58.2%) (p=0.000), and there was no significant difference in the proportion of patients who died within 28 days of ICU admission (p=0.943). Next, correlation coefficients of the independent variables were calculated for variable selection to avoid multicollinearity in the multiple regression analysis model (Table 5). The variables that were significantly correlated with ICU LOS were gender (r=0.07), ICU readmission status (r=0.34), and outcome (r=0.16).

**Table4.** The primary and secondary outcome measures in the before and after groups (N=1765)

| Outcome mesures | Bfore group<br>(n=898) | After group<br>(n=867) |  |
| --- | --- | --- | --- |
| ICU LOS, median day (IQR) | 3.0 (2.0 - 5.0) | 3.0 (2.0 - 5.0) | 0.859 |
| Hospital LOS, median day (IQR) | 20.0 (11.0 - 37.0) | 19.0 (10.0 - 31.0) | 0.023 |
| The proportion of patients who started nutrition within 48 hours of ICU admission, % | 61.9 | 67.8 | 0.009 |
| The proportion of patients who started rehabilitation within 48 hours of ICU admission, % | 58.2 | 68.0 | <0.001 |
| Mortality within 28 days of ICU admission, % | 7.2 | 7.1 | 0.943 |

The Mann-Whitney U test and $\chi^2$ test were used to compare primary and secondary outcome measures in the before and after the Introduction of interprofessional conferences.
Before group : the group before the introduction of conferences, After group : the group after the introduction of conferences
ICU LOS : intensive care unit length of stay, Hospital LOS : hospital length of stay, IQR : inter quartile range

**Table5.**
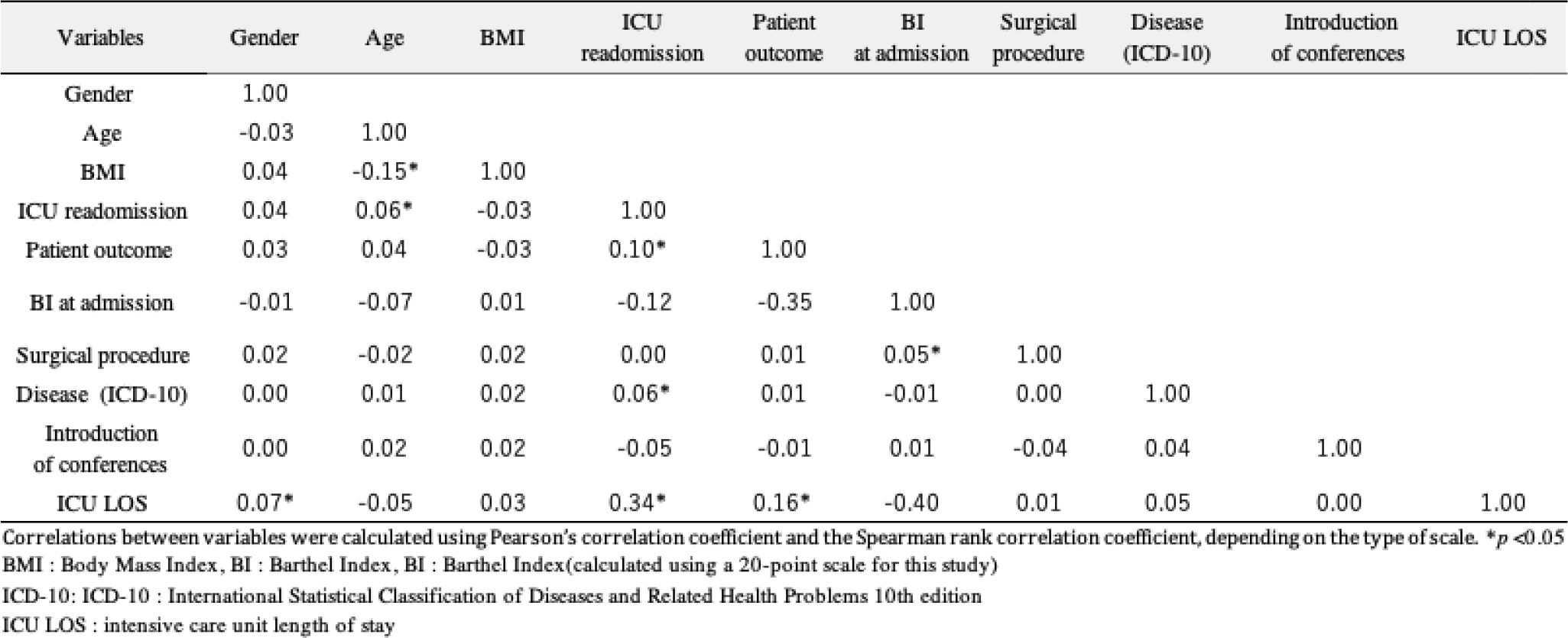
Correlation coefficients of the variables (N=1764)

Additionally, a Poisson regression analysis was performed to determine the association between the primary outcome (ICU LOS) and the introduction of interprofessional conferences (Table 6). The Poisson regression analysis revealed a significant association between ICU LOS and the introduction of interprofessional conferences (partial regression coefficient: -0.08, 95% confidence interval [CI]: -0.13 to -0.04; incidence rate ratio: 0.91, 95% confidence interval: 0.87 to 0.95). Other significant variables were gender (partial regression coefficient: 0.09; 95% CI: 0.04 to 0.13), age (partial regression coefficient:-0.01; 95% CI: -0.01 to 0.00), ICU readmission (partial regression coefficient:1.24; 95% CI: 1.18 to 1.29), and the following diseases: neoplasms (partial regression coefficient:-0.38; 95% CI: -0.44 to -0.32); gastrointestinal diseases (partial regression coefficient:-0.12; 95% CI: -0.19 to -0.05); and other diseases (partial regression coefficient:0.15; 95% CI: 0.09 to 0.20). A multiple regression analysis showed a significant association between hospital LOS and the introduction of interprofessional conferences (partial regression coefficient: -2.96; 95% CI: -5.20 to -0.72) (Table 7). There was also a significant association between whether nutrition was started within 48 hours of ICU admission and the introduction of interprofessional conferences (odds ratio: 1.45; 95% CI: 1.14 to 1.84). The introduction of interprofessional conferences was also significantly associated with whether rehabilitation was started within 48 hours of ICU admission (odds ratio: 0.77; 95% CI: 0.51 to 1.17). However, there was no significant association between the introduction of interprofessional conferences and mortality within 28 days of ICU admission (odds ratio: 0.77; 95% CI: 0.51 to 1.17).

**Table6.** The association between the ICU LOS and the introduction of interprofessional conferences (N=1724)

| Variables |  | Model1 (Crude) |  |  | Model1 (Ajusted 1) |  |  | Model1 (Ajusted 2) |  |  |
| --- | --- | --- | --- | --- | --- | --- | --- | --- | --- | --- |
|  |  | Regression coefficient | 95% CI |  | Regression coefficient | 95% CI |  | IRR | 95% CI |  |
|  |  |  | Lower bound | Upperbound |  | Lower bound | Upperbound |  | Lower bound | Upperbound |
| Interprofessional conferences | Bfore group | Reference |  |  | Reference |  |  | Reference |  |  |
|  | After group | -0.173 *** | -0.215 | -0.131 | -0.084 *** | -0.128 | -0.042 | 0.913 *** | 0.872 | 0.956 |
| Gender | Female | Reference |  |  | Reference |  |  | Reference |  |  |
|  | Male | 0.189 *** | 0.146 | 0.232 | 0.085 *** | 0.041 | 0.130 | 1.106 *** | 1.056 | 1.158 |
| Age |  | -0.001 | -0.002 | 0.000 | -0.005 *** | -0.007 | -0.004 | 0.995 *** | 0.994 | 0.997 |
| BMI |  | 0.005 ** | 0.000 | 0.010 | 0.004 | 0.000 | 0.009 | 1.015 *** | 1.009 | 1.020 |
| ICU readmission | Without | Reference |  |  | Reference |  |  | Reference |  |  |
|  | With | 1.402 *** | 1.348 | 1.456 | 1.235 *** | 1.180 | 1.292 | 2.965 *** | 1.180 | 1.292 |
| Patient outcome | Alive | Reference |  |  | Reference |  |  | Reference |  |  |
|  | Dead | 0.241 *** | 0.228 | 0.254 | 0.156 *** | 0.143 | 0.171 | 1.116 *** | 1.098 | 1.135 |
| BI at admission |  | -0.040 *** | -0.042 | -0.038 | -0.016 *** | -0.019 | -0.013 | 0.980 *** | 0.977 | 0.983 |
| Surgical procedure | Without | Reference |  |  | Reference |  |  | Reference |  |  |
|  | With | 0.000 | -0.051 | 0.051 | -0.003 | -0.055 | 0.049 | 1.067 ** | 1.009 | 1.130 |
| Disease (ICD-10) | Cardiovascular disease | Reference |  |  | Reference |  |  | Reference |  |  |
|  | Neoplasms | -0.511 *** | -0.566 | -0.457 | -0.377 *** | -0.436 | -0.319 | 0.948 | 0.895 | 1.004 |
|  | Gastrointestinal diseases | 0.136 ** | 0.068 | 0.205 | -0.116 ** | -0.188 | -0.046 | 0.973 | 0.982 | 1.117 |
|  | Other diseases | 0.273 *** | 0.219 | 0.327 | 0.146 *** | 0.089 | 0.204 | 1.047 | 0.899 | 1.053 |

Poisson regression analysis was used. Ajusted 1 is calculated by regression coefficient, and Ajusted 2 is calculated by IRR. R<sup>2</sup> = 0.241 for Ajusted 1, R<sup>2</sup> = 0.167 for Ajusted 2, \*\*p<0.01, \*\*\*p<0.001
95%CI : 95% confidence interval, IRR : Incidence rate-ratio, BMI : Body Mass Index, BI : Barthel Index(calculated using a 20-point scale for this study)
ICU LOS : intensive care unit length of stay, before group : the group before the introduction of conferences, after group : the group after the introduction of conferences
ICD-10 : International Statistical Classification of Diseases and Related Health Problems 10th edition

**Table7.**
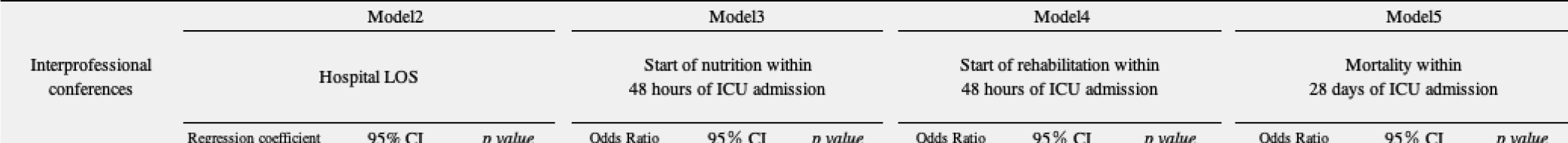
The association between secondary outcome measures and the introduction of interprofessional conferences (N=1724)

## Discussion

### Effectives of interprofessional conferences on starting nutrition and rehabilitation

The start of nutrition within 48 hours of ICU admission and the start of rehabilitation within 48 hours of ICU admission were significantly higher in the after group than in the before group. The benefits of interprofessional team care include the promotion of shared decision-making among various medical professionals, patients, and family members, the reduction of medical errors and disadvantages to patients, and the improvement of the quality of evidence-based care [3,5,17]. The Japanese guidelines for nutritional therapy for critically ill patients recommend early enteral nutrition within 24 to 48 hours after ICU admission [18]. During early rehabilitation, it is also recommended that positioning and early ambulation practices should be the basis for the prevention of respiratory complications with acute respiratory failure in the ICU [19]. Therefore, it is thought that interprofessional communication was promoted at the interprofessional conferences, and that the exchange of knowledge, skills, and experience among the various medical professionals led to the provision of care based on the evidence of previous studies, which led to the early start of nutrition and rehabilitation. Information sharing has been identified as one of the factors associated with interprofessional collaboration that influence early rehabilitation in the ICU [20]. Information sharing is important when discussing how each professional perceives the patient’s condition based on expertise and when discussing specific care among multiple professionals who are working together. Furthermore, it has been reported that increased information sharing opportunities have led to the improved ability to perform activities of daily living of patients in general surgical wards [21], and that facilitating information sharing among multiple professionals may promote early rehabilitation, prevent complications, and improve the long-term prognosis in the ICU [20]. Therefore, the introduction of interprofessional conferences may have increased opportunities for information sharing, thus leading to the shared recognition of the characteristics of each professional and patient, which may have contributed to early rehabilitation.

### Effects of interprofessional conferences on intensive care unit length of stay and hospital length of stay

Although there was no significant difference between the two groups according to the univariate analysis, adjusting for gender, age, BMI, ICU readmission, outcome, BI at admission, and disease (ICD-10) in the multivariate analysis resulted in significantly lower ICU LOS compared to that before the introduction of interprofessional conferences. Previous studies have described the possibility that interprofessional rounds, during which an interprofessional team performs daily rounds at the patient’s bedside, facilitate the implementation of respiratory therapy and nurse protocols for sedation, thus leading to shorter ventilator use and ICU LOS [22,23]. One study [24] reported that less than 10% of residents and nurses understood the goals of care; however, after the introduction of daily goal sheets and interprofessional sharing, the percentage of residents and nurses who understood the goals of care increased to more than 95%. As a result, there was a reduction in the ICU LOS. Based on previous studies, it can be inferred that sharing patient goals and the care and treatment provided by multiple professions contribute to the ICU LOS and outcomes [24]. During this study, one of the objectives of the interprofessional conferences was to share the care goals and patient goals; regardless of the format of the interprofessional rounds and conferences, the sharing of inpatient conditions and goals led to improved patient outcomes, such as shorter ICU LOS. Furthermore, after the introduction of interprofessional conferences, more patients started nutrition within 48 hours of admission to the ICU, and more patients started rehabilitation within 48 hours of admission to the ICU. Although robust evidence of early enteral nutrition is not available, a previous study reported that early initiation of nutrition improved hospital LOS and ICU LOS [25]. Additionally, early ambulation and early active exercise for patients admitted to the ICU have been shown to reduce ICU LOS [26] and hospital LOS [27–29]. Therefore, it is suggested that, in addition to shared patient goals among professionals, improved care processes, such as the early start of nutrition and rehabilitation, contributed to the reductions in ICU LOS and hospital LOS.

### Study implications

During this retrospective study, we were able to use a large amount of original data by extracting a sample from the medical records. This made it possible to compare our outcomes with those of other studies of interprofessional collaboration. This is the first study to examine the effectiveness of interprofessional conferences in the ICU after adjusting for confounding factors through a multivariate analysis. The results confirmed that shorter ICU stays, shorter hospital LOS, and the earlier start of nutrition and rehabilitation were achieved after the introduction of these conferences. Furthermore, the results suggested that if the common goals are to promote collaboration among multiple professionals, to activate communication between professionals, and to share patient conditions, goals, and care among professionals, then patient outcomes will improve regardless of the form of collaboration (such as interprofessional rounds, interprofessional protocols, and interprofessional conferences).

### Study limitations

There were three limitations to this study. First, it was a before-and-after comparative study involving patient data extracted from the medical records. Adjusting for confounding factors associated with changes in healthcare delivery systems and treatment methods over time was not possible. Additionally, there was no clear definition of the target patients involved in interprofessional conferences, which tended to focus on critically ill patients, patients whose treatment or care was not progressing as planned, or patients at high risk for such problems. Adjusting for confounding factors related to the patient status, such as disease severity, history of illness, and presence of delirium, was not possible. Therefore, future prospective studies with random sampling are required to examine the effectiveness of interprofessional conferences in more detail. Second, the study was conducted at a single center and the sample was biased, which affected the generalizability of the study results. Third, the study was not able to compare participants who attended the interprofessional conferences with those who did not. Only 22 patients were targeted by the conferences. Therefore, after accumulating more cases, it will be necessary to conduct a multivariate analysis to control for confounding factors and to clarify causal inferences based on propensity scores.

## Conclusion

After the introduction of interprofessional conferences in the ICU, more patients started nutrition within 48 hours of ICU admission, more patients started rehabilitation within 48 hours of ICU admission, and shorter ICU LOS and hospital LOS were observed.

## Data Availability

All relevant data are within the manuscript.

## Acknowledgments

We would like to express our sincere gratitude to all patients who provided their medical records for this study, to Mr. Ohnuma of the System Development Division of the Information Systems Department at the study facilities for his great cooperation in data collection, and to Associate Professor Uranaka and Associate Professor Asazawa for their guidance during this study.

